# Validation of the Kansas City Cardiomyopathy Questionnaire in Patients with Tricuspid Regurgitation: The Tri-QOL Study

**DOI:** 10.1101/2024.02.07.24302462

**Authors:** Suzanne V. Arnold, John A. Spertus, Kensey Gosch, Shannon M Dunlay, Danielle M. Olds, Philip G. Jones, Fraser D. Bocell, Changfu Wu, David J. Cohen

## Abstract

**Importance:** Improving patients’ health status is a key goal of treating tricuspid regurgitation (TR). The Kansas City Cardiomyopathy Questionnaire (KCCQ) is a heart-failure disease specific health status measure that has been used to capture the health status impact of TR and the benefit of transcatheter tricuspid valve intervention (TTVI), but its validity in this clinical setting is unknown.

**Objective:** To evaluate the psychometric properties of the KCCQ in patients with TR.

**Design, Setting, Participants:** Data were pooled from patients with severe TR enrolled in 11 manufacturer-sponsored trials of TTVI. The data were transferred to the FDA to harmonize and anonymize prior to analysis by an independent center. Prespecified analyses included evaluation of internal consistency, reproducibility, responsiveness, construct validity, and prognostic validity.

**Results:** The study cohort comprised 2693 patients (mean age 79±8 years, 62% women, mean baseline KCCQ Overall Summary [KCCQ-OS] score 50±23), enrolled in either single-arm (n=1517) or randomized (n=1176) investigations of TTVI. There was strong internal consistency within individual domains (Cronbach’s alpha 0.77-0.83). Among clinically stable patients between 1 and 6 months, there were small mean changes in KCCQ domain and summary scores (differences of -0.1 to 1.9 points), demonstrating reproducibility. In contrast, domain and summary scores of patients who underwent TTVI showed large improvements at 1-month after treatment (mean changes 12.1-21.4 points), indicating excellent responsiveness. Construct validity was moderately-strong to strong when the domains were compared with best available reference measures (Spearman correlations 0.47-0.69). The KCCQ-OS was strongly associated with clinical events, with lower scores being associated with an increased risk of mortality (HR 1.34 per 10-point decrement, 95% CI 1.22-1.47) and heart failure hospitalization (HR 1.24 per 10-point decrement, 95% CI 1.17-1.31).

**Conclusions and Relevance:** The KCCQ has strong psychometric properties, including reliability, responsiveness, and validity in patients with severe TR. These data support its use in clinical investigations of TTVI as a measure of the impact of treatment on patients’ symptoms, function, and quality of life.

**KEY POINTS:** *Question:* Is the Kansas City Cardiomyopathy Questionnaire (KCCQ) a valid disease-specific health status measure in patients with severe tricuspid regurgitation (TR)?

*Findings:* Data on 2693 patients from 11 clinical trials of various transcatheter tricuspid valve interventions (TTVI) from 2 device manufacturers were pooled and then analyzed by an academic analytic center. We found that the KCCQ has strong psychometric properties, including reliability, responsiveness, and validity in patients with severe TR.

*Meaning:* These data support continued use of the KCCQ in clinical trials of TTVI as a measure of the impact of treatment on patients’ symptoms, function, and quality of life.

---

Beyond its impact on survival, severe tricuspid regurgitation (TR) is associated with symptoms of dyspnea, fatigue, and edema that result in reduced functional capacity^1,2^ and impaired quality of life.^3–9^ ^15–17^ In recent years, a number of transcatheter tricuspid valve interventions (TTVI) have been introduced,^2,18^ which are less invasive than cardiac surgery and may increase the number of patients potentially eligible for TR treatment. Since improving health status is a key treatment goal for patients with TR, it is critically important to reliably, validly, and sensitively measure patient-reported health status to quantify the benefit of TTVI from patients’ perspectives. The Kansas City Cardiomyopathy Questionnaire (KCCQ) is a heart-failure specific health status questionnaire that was originally developed for use in patients with heart failure with reduced left ventricular ejection fraction.^19^ It has subsequently undergone extensive reliability and validity testing in other forms of left-sided heart failure^20–24^ and was used as the primary health status outcome in most of the pivotal clinical trials of transcatheter aortic and mitral valve interventions.^25–32^ It has also been used as the primary disease-specific health status outcome for many clinical investigations of TTVI,^3–5,8,33,34^ but its validity and psychometric properties in this clinical setting are unknown. Given the prominent role of health status in the evaluation of TTVI, we used data from multiple clinical trials to assess the reliability, responsiveness, validity, and prognostic importance of the KCCQ among patients with TR.

## METHODS

### Study Design

The TRI-QOL study is an investigator-initiated study funded by an FDA contract to examine the validity of the KCCQ in patients with TR. As a part of this study, we collaborated with the device manufacturers to obtain patient-level data from multiple TTVI trials (Supplemental Table 1), including 8 single-arm and 3 randomized trials of various TTVI conducted by Edwards Lifesciences (Irvine, CA) or Abbott (Santa Clara, CA). Patient-level data were transferred to the FDA to harmonize and anonymize the variables and then transferred to the academic analytic center for this study. The analytic plan was developed by the investigators and executed independently of the device manufacturers that contributed the data, and there was no formal or informal review of the results or manuscript by the device manufacturers prior to submission. The investigators were blinded to the trial sponsors, the devices under study, and, in randomized trials, to treatment assignment. Each trial received appropriate review and approval for initial data collection, including written informed consent from all participants. These secondary psychometric analyses were approved by the Advarra central institutional review board, and a waiver of informed consent was granted, given the anonymized nature of the data transferred to the analytic center.

### Health Status and Clinical Assessment

The KCCQ is a 23-item self-administered questionnaire that addresses specific health domains pertaining to heart failure: physical limitations, symptoms, quality of life, social limitation, symptom stability, and self-efficacy.^19^ The physical limitations and symptoms domains are combined into a clinical summary score (KCCQ-CS), with the quality of life and social limitations domains added to form the overall summary score (KCCQ-OS). Values for each domain and the summary scores range from 0 to 100 with higher scores indicating lower symptom burden and better quality of life. The symptom stability domain uses a single question to assess recent changes in patients’ heart failure symptoms, is not appropriate for internal consistency or longitudinal analyses, and was excluded. The self-efficacy domain is designed to assess whether a patient feels they have the knowledge and skills to manage their heart failure and is not a direct measure of patients’ health status, so it was also excluded from these analyses. As such, our analyses focused on the clinical and overall summary scores and the four domains that contribute to them.

As additional measures to explore the validity of the KCCQ, generic health status was assessed with the Medical Outcomes Study Short-Form 12 (SF-12) Health Survey,^25^ a reliable and valid measure of generic health status that provides summary component scores for overall physical (PCS) and mental (MCS) health.^35^ Scores are standardized using norm-based methods to generate a mean of 50 and a standard deviation of 10, with higher scores indicating better health status.^36^ Data were also collected on physician-estimated New York Heart Association (NYHA) class and 6-minute walk test distance. The included trials had different time points of health status assessment and follow-up, and thus not all patients were included in each analysis.

### Statistical Analysis

Since the identity of the specific trials was masked, analyses were performed among all eligible patients without adjustment for treatment, unless otherwise specified. Where appropriate, analyses were performed exclusively in the single-arm studies, in which all patients underwent TTVI. The specific analytic cohort and the tests used to assess each of the psychometric properties are described in detail below and summarized in Supplemental Table 2. All analyses were repeated after stratification by sex of the participant, to explore whether there were any meaningful differences in the psychometric properties of the KCCQ by sex. Analyses were conducted using SAS v9.2 (SAS Institute, Inc., Cary, NC), and statistical significance was determined by a 2-sided p-value of <0.05.

#### Determining Questionnaire Reliability

Internal consistency of the KCCQ domains at baseline was assessed among all patients using Cronbach’s alpha, which ranges from 0-1 and reflects the internal consistency of different items within each individual domain. An α≥0.9 indicates excellent consistency (but may also indicate redundancy); 0.9>α≥0.8 good consistency; 0.8>α≥0.7 acceptable consistency; 0.7>α≥0.6 questionable consistency; and α<0.6 poor consistency.^37^ For test-retest reproducibility, we identified a clinically-stable cohort between two timepoints—patients who were alive at 6 months and had no change in NYHA class and no heart failure hospitalizations between 1 and 6 months. Within this cohort, we calculated the mean change in each KCCQ domain and summary score between 1 and 6 months. Reproducibility was then tested using the intraclass correlation coefficient (ICC),^38^ which is the ratio of between-group variance to total variance and ranges from 0 to 1, with higher scores indicating greater test-retest reproducibility (0-0.2 indicates poor agreement, 0.3-0.4 fair agreement, 0.5-0.6 moderate agreement, 0.7-0.8 strong agreement, and >0.8 excellent agreement).^39^

#### Determining Questionnaire Responsiveness

The responsiveness of the KCCQ domains to a clinical change was assessed among patients from the single-arm studies who underwent TTVI and were alive 1 month after their procedure. Scores at baseline and 1 month were compared using paired *t*-tests. Cohen’s *d* effect size,^40^ which quantifies the magnitude of change relative to baseline variation, was also used to assess the responsiveness of the questionnaire to clinical change. An effect size of 0.2 to 0.3 indicates a small effect; 0.5 a moderately large effect; and ≥0.8 a large effect.^41^

#### Determining Questionnaire Validity

The validity of the different domain and summary scores was evaluated by Spearman’s correlations (and associated 95% confidence interval [CI]) between the scores and other prespecified measures that quantify similar concepts. For these analyses, the analytic cohort consisted of all patients alive at 1 month to allow for a wide range of KCCQ scores (including patients who were managed medically or with TTVI). For the physical limitations domain, we examined correlations with the 6-minute walk test, SF-12 PCS, and NYHA class. For the symptoms domain, KCCQ-CS, and KCCS-OS, we examined their correlations with NYHA class. Finally, we examined the correlations between the quality of life and social limitations scales with the SF-12 MCS. The criterion standards chosen for these comparisons closely mirror those used in the original validation of the KCCQ.^19^

To examine the ability of the KCCQ to discriminate between different levels of change, we calculated the change in domain and summary scores for all patients from baseline to 1 month stratified by their concurrent change in NYHA class, comparing these using a linear trend tests. Finally, among patients in the single-arm studies, we used linear trend tests to compare the change in each KCCQ domain and summary score from baseline to 1 month after TTVI with the change in TR grade (assessed by core laboratories on a 6-level scale^42^). For these analyses, we excluded patients who were treated with the FORMA device, owing to challenges in determining follow-up TR grade with a spacer device.^18^ We also constructed linear regression models for change in KCCQ (domain and summary scores) from baseline to 1 month as a function of change in TR grade, adjusted for baseline TR grade and baseline KCCQ-OS score (modeled using restricted cubic splines).

#### Prognostic Importance

We examined two questions about prognosis with the KCCQ in patients with TR: 1) the association of KCCQ-OS score with subsequent risk of outcomes, and 2) the association of a change in KCCQ-OS after TTVI with subsequent risk of outcomes. The first analysis used all patients at 1 month (including control patients from the randomized trials), in order to provide a wide range of KCCQ scores. We first examined the relationship between KCCQ-OS at 1 month as a categorical variable (0-25, 26-50, 51-75, and >75)^21^ and 1-year death, heart failure hospitalization, and the composite of death or heart failure hospitalization using Kaplan-Meier event curves. Cox proportional hazards regression was used to evaluate the independent association between 1-month KCCQ-OS as a continuous linear variable (restricted cubic splines were not significant) and 1-year death, heart failure hospitalization (censored at death), and death or heart failure hospitalization. Models were adjusted for age, sex, body mass index, chronic lung disease, atrial fibrillation/flutter, coronary artery disease, prior myocardial infarction, prior coronary artery bypass graft surgery, prior stroke, permanent pacemaker, and left ventricular ejection fraction. Continuous covariates were modeled using restricted cubic splines to accommodate potential nonlinear associations.

We restricted the second analysis to patients from the single-arm trials who had undergone TTVI, to specifically examine the prognostic importance of changes in KCCQ after intervention. We used Cox proportional hazards regression models to test the association of change in KCCQ-OS at 1 month as a continuous linear variable (restricted cubic splines were not significant) with 1-year death, heart failure hospitalization (censored at death), and death or heart failure hospitalization. These models adjusted for baseline KCCQ-OS (modeled non-linearly) and the patient covariates listed above.

## RESULTS

### Patient Population

The analytic cohort comprised 2693 patients with symptomatic TR, which was severe in 34%, massive in 29%, and torrential in 31% (Table 1). Mean age of the cohort was 78.6±8.0 years, 62% were female, 92% had atrial fibrillation or flutter, and 63% were enrolled in the United States. Mean baseline KCCQ-OS was 50.3±22.8.

**Table 1.** Baseline characteristics of study cohort.

|  |  |
| --- | --- |
| Enrolled in United States | 62.6% (1686/2693) |
| Enrolled in randomized trial | 43.7% (1176/2693) |
| Age, years | 78.6 ± 8.0 (2693) |
| Women | 61.6% (1658/2693) |
| Body mass index, kg/m<sup>2</sup> | 26.6 ± 5.6 (2685) |
| Hypertension | 84.2% (1345/1597) |
| Pulmonary hypertension | 63.2% (947/1498) |
| Chronic lung disease | 15.2% (405/2668) |
| Atrial fibrillation/flutter | 91.5% (2464/2693) |
| Coronary artery disease | 18.0% (485/2693) |
| Prior myocardial infarction | 9.8% (264/2693) |
| Prior percutaneous coronary intervention | 19.2% (517/2693) |
| Prior coronary bypass graft surgery | 15.7% (424/2693) |
| Prior aortic valve surgery | 15.8% (340/2152) |
| Prior mitral valve surgery | 23.2% (500/2152) |
| Pacemaker or implantable defibrillator | 25.8% (689/2668) |
| Prior stroke | 10.0% (269/2693) |
| Heart failure hospitalization in the past 12 months | 38.5% (1035/2687) |
| Tricuspid regurgitation grade |  |
| None/trace/mild | 0.3% (8/2569) |
| Moderate | 5.8% (148/2569) |
| Severe | 33.9% (872/2569) |
| Massive | 29.2% (749/2569) |
| Torrential | 30.8% (792/2569) |
| Left ventricular ejection fraction, % | 55.8 ± 10.3 (2302) |
| TAPSE, cm | 1.6 ± 0.4 (2328) |
| Right ventricular FAC, % | 37.9 ± 8.1 (2198) |
| Kansas City Cardiomyopathy Questionnaire Scores |  |
| Physical Limitation | 53.1 ± 25.5 (2613) |
| Symptoms | 54.5 ± 25.1 (2643) |
| Quality of Life | 45.1 ± 24.6 (2643) |
| Social Limitation | 48.3 ± 30.1 (2532) |
| Clinical Summary | 53.8 ± 23.1 (2645) |
| Overall Summary | 50.3 ± 22.8 (2645) |
| New York Heart Association class |  |
| 1 | 0.6% (16/2687) |
| 2 | 28.5% (765/2687) |
| 3 | 65.7% (1765/2687) |
| 4 | 5.2% (141/2687) |

TAPSE, tricuspid annular plane systolic exertion (normal >1.7 cm); FAC, fractional area of change (normal >35%)

### Reliability of the KCCQ

The quality of life and social limitations domains demonstrated acceptable internal consistency (Cronbach’s alpha=0.77 and 0.78, respectively) while all other domains showed good or excellent consistency (Table 2). Among 803 patients who were deemed clinically stable, the mean differences between 1-and 6-month KCCQ scores were <1 point for all domain and summary scores, except the quality of life domain for which the mean within-patient change was +1.9 points (95% CI 0.4 to 3.4; Table 2). The ICC was moderate-to-high for all domain and summary scores (range 0.63-0.71) demonstrating that between-patient variability was greater than within-patient variability.

**Table 2.** Internal Consistency, Test-Retest Reproducibility, and Responsiveness of the KCCQ.

| **Property** | **Consistency** | **Test-Retest Reproducibility** |  | **Responsiveness** |  |
| --- | --- | --- | --- | --- | --- |
| **Analytic Cohort** | **All patients; n=2645** | **Stable patients; n=803** |  | **TTVI patients; n=1323** |  |
| **Time Frame** | **Baseline** | **1 to 6 Months** |  | **Baseline to 1 Month** |  |
| **Test** | **Cronbach's Alpha** | **Mean Difference**<br>**(95% CI)** | **ICC** | **Mean Difference**<br>**(95% CI)** | **Cohen's D** |
| KCCQ Domain Scores |  |  |  |  |  |
| Physical limitation | 0.83 | -0.1 (-1.4 to 1.3) | 0.68 | 12.1 (10.8 to 13.4) | 0.47 |
| Symptoms | 0.83 | 0.4 (-0.8 to 1.7) | 0.65 | 17.8 (16.5 to 19.1) | 0.71 |
| Quality of life | 0.77 | 1.9 (0.4 to 3.4) | 0.63 | 21.4 (20.0 to 22.8) | 0.88 |
| Social limitation | 0.78 | 0.9 (-0.9 to 2.7) | 0.63 | 16.6 (14.9 to 18.3) | 0.56 |
| KCCQ Summary Scores |  |  |  |  |  |
| Clinical summary | NA | 0.4 (-0.8 to 1.5) | 0.70 | 15.1 (13.9 to 16.2) | 0.65 |
| Overall summary | NA | 0.9 (-0.2 to 2.1) | 0.71 | 17.0 (15.9 to 18.2) | 0.75 |

KCCQ, Kansas City Cardiomyopathy Questionnaire; TTVI, transcatheter tricuspid valve intervention; Cronbach's Alpha ( $\alpha \geq 0.9$ indicates excellent consistency [but may also indicate redundancy]; $0.9 > \alpha \geq 0.8$ is good; $0.8 > \alpha \geq 0.7$ is acceptable; $0.7 > \alpha \geq 0.6$ is questionable; $\alpha < 0.6$ is poor); ICC, intra-class correlation (agreement: 0-0.2 poor, 0.3-0.4 fair, 0.5-0.6 moderate, 0.7-0.8 strong, $> 0.8$ excellent); Cohen's D (0.2 to 0.3 small effect; $\sim 0.5$ medium effect; $\geq 0.8$ large effect); NA, not applicable

### Responsiveness of the KCCQ

Among 1323 patients who underwent TTVI, there were moderate-to-large differences in the KCCQ domain and summary scores before and after intervention (Table 2). The mean KCCQ-OS increased from 49.8±22.6 before intervention to 66.9±22.3 1 month after intervention, with a mean difference of 17.0 points (95% CI 15.9 to 18.2) and an effect size (ratio of mean change to standard deviation at baseline) of 0.75. The physical limitations domain had the smallest change from baseline to 1 month (mean difference 12.1 points, effect size 0.47), while the quality of life domain demonstrated the largest change (mean difference 21.4 points, effect size 0.88).

### Criterion Validity of the KCCQ

The KCCQ domain and summary scores demonstrated moderately-strong to strong correlations with the best available reference measures (Spearman correlations 0.47-0.69; Table 3). When comparing change in KCCQ versus concurrent change in NYHA class from baseline to 1 month, there were strong linear correlations between the two measures, with substantially larger increases in both domain and summary scores among those with greater reductions in NYHA class (Table 4). Among patients who underwent TTVI, there was also a graded association between the change in KCCQ scores from baseline to 1 month and concurrent change in TR grade (Table 5). Finally, in regression models that adjusted for baseline KCCQ and baseline TR grade, there were strong, independent associations between change in TR grade from baseline to 1 month and concurrent changes in all domain and summary scores, with an estimated increase of 2.7 points on the KCCQ-OS (95% CI 1.5 to 3.9) for each 1-grade reduction in TR (Table 5).

**Table 3.**
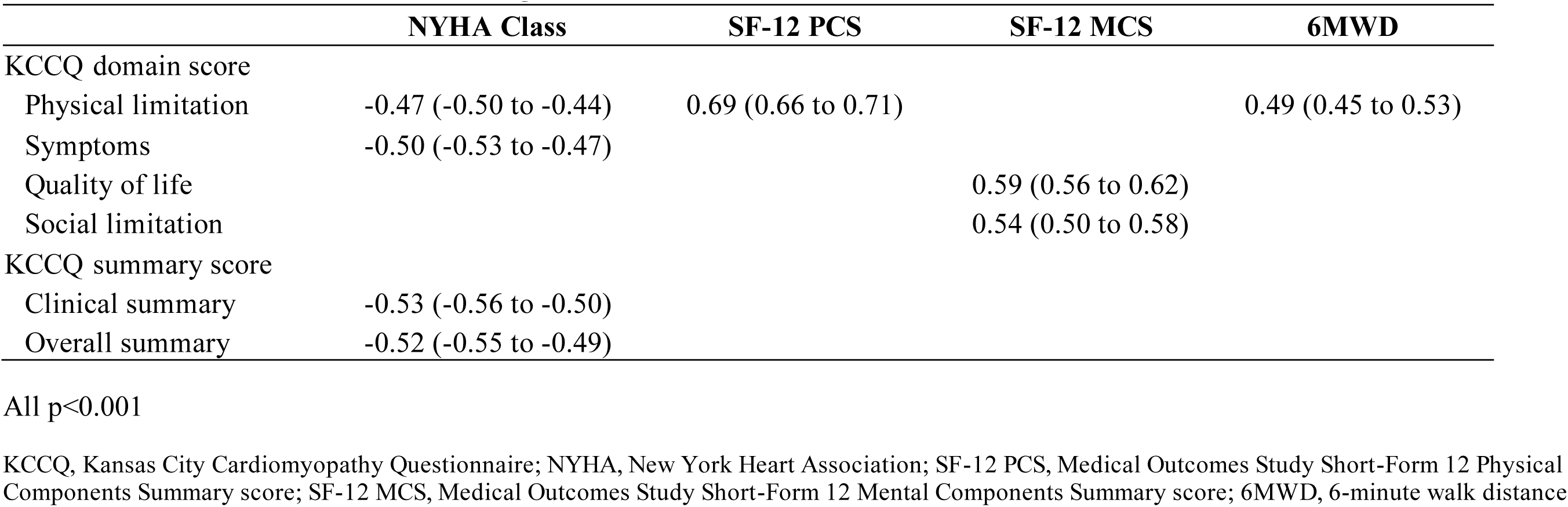
Spearman Correlations of KCCQ Domain and Summary Scores with Other Measures.

|  | **NYHA Class** | **SF-12 PCS** | **SF-12 MCS** | **6MWD** |
| --- | --- | --- | --- | --- |
| KCCQ domain score |  |  |  |  |
| Physical limitation | -0.47 (-0.50 to -0.44) | 0.69 (0.66 to 0.71) |  | 0.49 (0.45 to 0.53) |
| Symptoms | -0.50 (-0.53 to -0.47) |  |  |  |
| Quality of life |  |  | 0.59 (0.56 to 0.62) |  |
| Social limitation |  |  | 0.54 (0.50 to 0.58) |  |
| KCCQ summary score |  |  |  |  |
| Clinical summary | -0.53 (-0.56 to -0.50) |  |  |  |
| Overall summary | -0.52 (-0.55 to -0.49) |  |  |  |

All $p < 0.001$
KCCQ, Kansas City Cardiomyopathy Questionnaire; NYHA, New York Heart Association; SF-12 PCS, Medical Outcomes Study Short-Form 12 Physical Components Summary score; SF-12 MCS, Medical Outcomes Study Short-Form 12 Mental Components Summary score; 6MWD, 6-minute walk distance

**Table 4.** Mean Change in KCCQ Scores According to Change in NYHA Class from Baseline to 1 Month.

|  | **Change in NYHA Class from Baseline to 1 Month** |  |  |  | **p-value for trend** |
| --- | --- | --- | --- | --- | --- |
|  | **Improvement by 2 or 3**<br>**(n=340)** | **Improvement by 1**<br>**(n=1046)** | **No change**<br>**(n=837)** | **Worsening**<br>**(n=103)** |  |
| Physical limitation | 20.1 ± 24.6 | 13.8 ± 22.8 | 5.3 ± 21.5 | -6.1 ± 21.7 | <0.001 |
| Symptoms | 29.3 ± 23.2 | 18.0 ± 22.4 | 8.5 ± 22.3 | 0.9 ± 24.5 | <0.001 |
| Quality of life | 33.9 ± 26.3 | 22.7 ± 25.3 | 13.0 ± 24.9 | -1.1 ± 22.7 | <0.001 |
| Social limitation | 30.1 ± 31.2 | 16.6 ± 29.3 | 8.8 ± 28.6 | -5.2 ± 28.3 | <0.001 |
| Clinical summary | 24.7 ± 20.8 | 16.1 ± 19.8 | 7.0 ± 19.3 | -2.8 ± 21.2 | <0.001 |
| Overall summary | 28.3 ± 21.3 | 18.0 ± 20.6 | 9.0 ± 20.0 | -2.8 ± 20.5 | <0.001 |

KCCQ, Kansas City Cardiomyopathy Questionnaire; NYHA, New York Heart Association

**Table 5.** Mean Change in KCCQ Scores According to Change in TR Grade at 1 Month after TTVI.

|  | Change in TR Grade from Baseline to 1 Month |  |  |  |  | p-value | Estimate (95% CI) for<br>change in KCCQ per 1-grade reduction in TR<sup>a</sup> | p-value |
| --- | --- | --- | --- | --- | --- | --- | --- | --- |
|  | -4<br>(n=109) | -3<br>(n=299) | -2<br>(n=415) | -1<br>(n=268) | 0<br>(n=108) |  |  |  |
| Physical limitation | 13.2 ± 24.1 | 16.0 ± 23.9 | 11.7 ± 23.8 | 8.7 ± 24.6 | 8.3 ± 19.2 | 0.012 | 1.5 (0.1 to 3.0) | 0.035 |
| Symptoms | 20.8 ± 24.8 | 21.2 ± 23.5 | 18.1 ± 22.2 | 13.5 ± 23.1 | 11.0 ± 21.9 | <0.001 | 2.9 (1.7 to 4.2) | <0.001 |
| Quality of life | 24.6 ± 29.9 | 25.1 ± 25.9 | 22.8 ± 25.8 | 17.9 ± 25.3 | 12.7 ± 23.5 | <0.001 | 3.4 (2.0 to 4.9) | <0.001 |
| Social limitation | 23.2 ± 32.2 | 20.6 ± 30.7 | 16.3 ± 29.5 | 11.9 ± 28.5 | 10.2 ± 25.8 | <0.001 | 2.8 (1.0 to 4.6) | 0.002 |
| Clinical summary | 17.2 ± 22.6 | 18.8 ± 20.9 | 15.1 ± 20.0 | 11.1 ± 20.6 | 9.6 ± 17.2 | <0.001 | 2.3 (1.2 to 3.5) | <0.001 |
| Overall summary | 20.3 ± 24.7 | 20.7 ± 21.3 | 17.4 ± 20.8 | 13.5 ± 20.8 | 10.3 ± 18.3 | <0.001 | 2.7 (1.5 to 3.9) | <0.001 |

KCCQ, Kansas City Cardiomyopathy Questionnaire; TR, tricuspid regurgitation; TTVI, transcatheter tricuspid valve intervention
<sup>a</sup> Adjusted for baseline KCCQ-OS and baseline TR grade

### Prognostic Importance

Among 2345 patients with 1-month assessments, there was a graded relationship between KCCQ-OS at 1 month and the incidence of death, heart failure hospitalization, and the composite of death or heart failure hospitalization through 1 year of follow-up (Figure 2A-C; Table 6). In Cox proportional hazards models, every 10-point decrement in the KCCQ-OS score was associated with an HR of 1.34 (95% CI 1.22-1.47) for death, 1.24 (95% CI 1.17-1.31) for heart failure hospitalization, and 1.27 (95% CI 1.20-1.33) for the composite of death or heart failure hospitalization (Table 6). Among 1323 patients in the single-arm trials who underwent TTVI, changes in KCCQ-OS at 1 month after intervention were also strongly associated with the risk of subsequent outcomes. In Cox proportional hazards models adjusted for baseline KCCQ-OS, every 10-point improvement in the KCCQ-OS score at 1 month was associated with an HR of 0.78 (95% CI 0.68-0.89) for death, 0.79 (95% CI 0.72-0.87) for heart failure hospitalization, and 0.79 (95% CI 0.73-0.85) for the composite of death or heart failure hospitalization (Table 6). In both analyses, the associations between KCCQ and subsequent outcomes were essentially unchanged after adjusting for patient demographic and clinical factors.

**Table 6.** Association of KCCQ-OS Score and Change in KCCQ-OS Score with Death and Heart Failure Hospitalization.

|  | Association of KCCQ-OS with Endpoint<sup>a</sup><br>(per 10-point decrement) |  |  |  | Association of Change in KCCQ-OS with Endpoint<sup>b</sup><br>(per 10-point increase) |  |  |  |
| --- | --- | --- | --- | --- | --- | --- | --- | --- |
|  | Unadjusted HR<br>(95% CI) | p-value | Adjusted HR<sup>c</sup><br>(95% CI) | p-value | Unadjusted HR<sup>d</sup><br>(95% CI) | p-value | Adjusted HR<sup>e</sup><br>(95% CI) | p-value |
| Death | 1.33 (1.21-1.46) | <0.001 | 1.34 (1.22-1.47) | <0.001 | 0.78 (0.68-0.89) | <0.001 | 0.81 (0.70-0.93) | 0.002 |
| HFH | 1.25 (1.18-1.32) | <0.001 | 1.24 (1.17-1.31) | <0.001 | 0.79 (0.72-0.87) | <0.001 | 0.80 (0.73-0.86) | <0.001 |
| Death or HFH | 1.27 (1.20-1.33) | <0.001 | 1.26 (1.19-1.32) | <0.001 | 0.79 (0.73-0.85) | <0.001 | 0.81 (0.74-0.87) | <0.001 |

KCCQ-OS, Kansas City Cardiomyopathy Questionnaire-overall summary score; HR, hazard ratio; CI, confidence interval; HFH, heart failure hospitalization
<sup>a</sup> KCCQ-OS at 1 month from trial enrollment, with endpoints assessed from 1 month through 1 year of follow-up.
<sup>b</sup> Change in KCCQ-OS from baseline to 1 month from TTVI, with endpoints assessed from 1 month through 1 year of follow-up; includes only patients in single-arm trials who were treated with TTVI.
<sup>c</sup> Adjusted for age, sex, body mass index, chronic lung disease, atrial fibrillation/flutter, coronary artery disease, prior myocardial infarction, prior coronary artery bypass graft surgery, prior stroke, permanent pacemaker, and left ventricular ejection fraction
<sup>d</sup> Adjusted for baseline KCCQ-OS
<sup>e</sup> Adjusted for baseline KCCQ-OS, age, sex, body mass index, chronic lung disease, atrial fibrillation/flutter, coronary artery disease, prior myocardial infarction, prior coronary artery bypass graft surgery, prior stroke, permanent pacemaker, and left ventricular ejection fraction

### Stratification by Sex

The demographic and clinical characteristics of the 1035 men and 1658 women are shown in Supplemental Table 3. Compared with men, women reported lower baseline KCCQ scores by ∼5-7-points across the domains and summary scores. Internal consistency, reproducibility, and responsiveness were similar in men and women (Supplemental Table 4), as were correlations of the domain and summary scores with available reference measures (Supplemental Table 5). The associations between change in KCCQ and change in NYHA class (Supplemental Table 6), change in KCCQ and change in TR grade (Supplemental Table 7), 1-month KCCQ-OS with subsequent death and heart failure (Supplemental Table 8), and change in KCCQ-OS at 1-month after TTVI with subsequent death and heart failure (Supplemental Table 8) were consistent between women and men. The one notable difference by sex was that the KCCQ appeared to be more responsive to clinical change in women versus men, with consistently higher mean changes and effect sizes across the domains and summary scores after TTVI—a finding that may reflect the lower KCCQ scores at baseline in women.

## DISCUSSION

With the development and refinement of transcatheter interventions to treat valvular heart disease, there has been increased recognition of the critical need to understand the impact of these innovative therapies on the symptoms, functional limitations, and quality of life of patients. As such, disease-specific health status, measured using the KCCQ, has been incorporated as a key secondary outcome for nearly all of the pivotal trials of transcatheter aortic valve replacement and transcatheter mitral valve repair and replacement therapies.^25–32^ As the field of transcatheter valvular interventions has expanded into treatment of TR, trials have continued to rely on the KCCQ to quantify the health status benefit of TTVI^3–5,8,33,34^—notwithstanding lack of evidence regarding the validity of this approach. While the KCCQ has been validated in a multiple forms of left-sided heart disease (including aortic stenosis),^19–24^ its performance as a disease-specific health status measure was unknown among patients with TR, where symptoms of right-sided heart failure may be more prominent. Given the importance of the KCCQ in defining the clinical benefit of TTVI, understanding the validity of the KCCQ in this clinical setting is even more critical.

In this study of >2500 patients with symptomatic TR enrolled in TTVI trials, we found that the KCCQ demonstrated similar performance as in patients with heart failure with reduced ejection fraction (HFrEF).^19^ The one difference compared with the original validation studies was in reproducibility, where ICCs in patients with TR for the domain and summary scores ranged from 0.63-0.71 (as compared with 0.82-0.92 in patients with HFrEF).^43^ These differences may be explained by the fact that our validation analyses were performed over a longer time frame for follow-up (5 months vs. 6 weeks) and without an additional criterion for clinical stability (patient and physician global health assessments) to ensure the identification of a stable cohort for test-retest reproducibility. The psychometric properties of the KCCQ in patients with TR were similar to those that were seen in patients with severe aortic stenosis.^23^ Finally, we found that KCCQ scores and changes in scores were strongly associated with the risk of subsequent death and heart failure hospitalization. Worse health status was associated with a higher risk of adverse outcomes, but improvement in health status after TTVI was associated with a reduction in that risk. The magnitude of these prognostic associations is similar to what has been observed in patients with chronic heart failure^21,44^ and after other transcatheter valve interventions.^45^

Importantly, we found similar performance of the KCCQ in men and women with TR. Women with heart failure generally report worse health status compared with men^46^—a difference also observed in our study. As a result, there have been concerns that men and women may interpret questions about symptoms, function, and quality of life differently. Prior quantitative^47^ and qualitative^48^ studies in patients with heart failure have generally dispelled this concern, but this was also essential to investigate in the current study, given the predominance of women in TTVI trials.

### KCCQ as a Clinical Trial Endpoint

Within TTVI trials, health status outcomes have played a particularly prominent role. Unlike transcatheter treatment of other valve conditions,^49,50^ TTVI has not yet been shown to reduce mortality or heart failure hospitalizations,^34^ making improvement in health status the key benefit of these procedures. As such, it was imperative to establish the validity of the KCCQ in this clinical setting. Although the KCCQ was not originally developed to describe the health status of patients with symptomatic TR, our analyses support the continued use of the KCCQ as a reliable and valid endpoint in TTVI trials.

In addition to demonstrating the validity of the KCCQ as a health status measure for patients with TR, several aspects of our study demonstrate the clinical relevance of the KCCQ for this population. First, we found a strong, independent correlation between the change in TR severity across a broad range of devices and short-term improvement in the KCCQ-OS score. Although these findings were based on open-label trials, the “dose-response” relationship among patients who all underwent device therapy is consistent with a true biologic effect of TR reduction on disease-specific health status. Moreover, we found strong associations between improvement in KCCQ-OS at 1 month after TTVI with a reduction in the risk of death and heart failure hospitalization (hazard ratios ∼0.8 per 10-point increase in KCCQ-OS), which provides further reassurance that differences in disease-specific health status are clinically meaningful. Long-term follow-up of ongoing randomized trials will be important to verify whether these expected benefits accrue to patients treated with various forms of TTVI.

### Limitations

Our study should be interpreted in the context of several potential limitations. First, not all KCCQ domains had independent reference standards for the validity analyses, so some domains were compared against the same metrics (e.g., the quality of life and social limitations domains were both compared against the SF-12 MCS), and theses metrics do not directly measure the same concepts of quality of life and social limitations, making a lower correlation to be expected, rather than a concern about the criterion validity of these scales. Second, although our analyses demonstrate strong psychometric performance of the KCCQ in patients with TR, we could not assess its content validity in the current study, such that there could be manifestations of TR not captured by KCCQ. For example, the KCCQ does not capture abdominal distension or anorexia, which may be prominent symptoms for some patients with symptomatic TR. Further, ongoing, qualitative work is needed to identify whether there are unique manifestations of TR that are not captured by the KCCQ. Finally, since these studies were not blinded to treatment, some assessments, such as NYHA class, may have been biased.

## Conclusions

In summary, in a study of >2500 patients enrolled in clinical trials of several forms of TTVI, the KCCQ was found to be a reliable, responsive, and valid disease-specific health status instrument for patients with TR. These results support its use as a clinically-relevant and patient-centered outcome measure for studies of TTVI.

**Figure 1.**
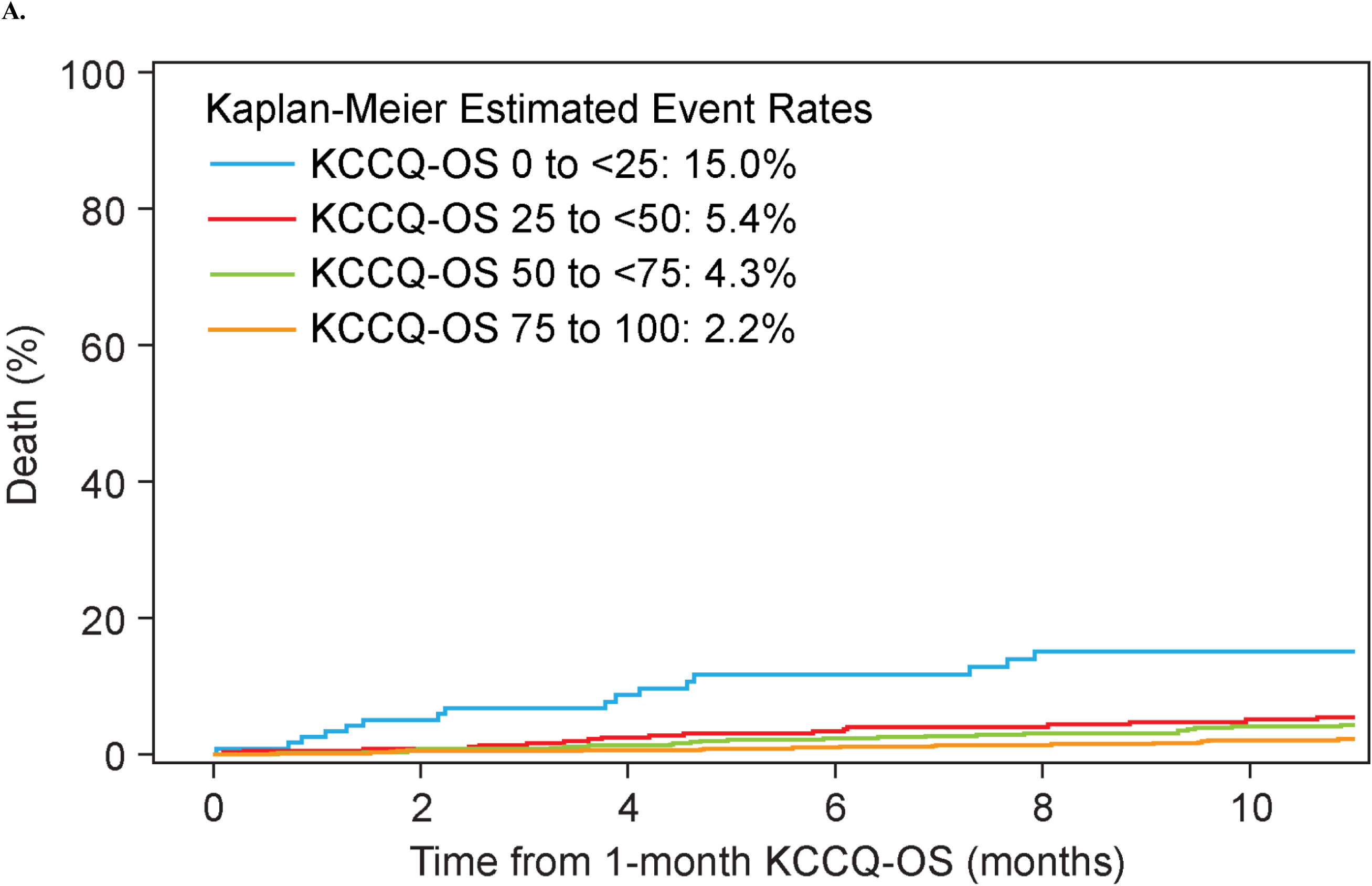
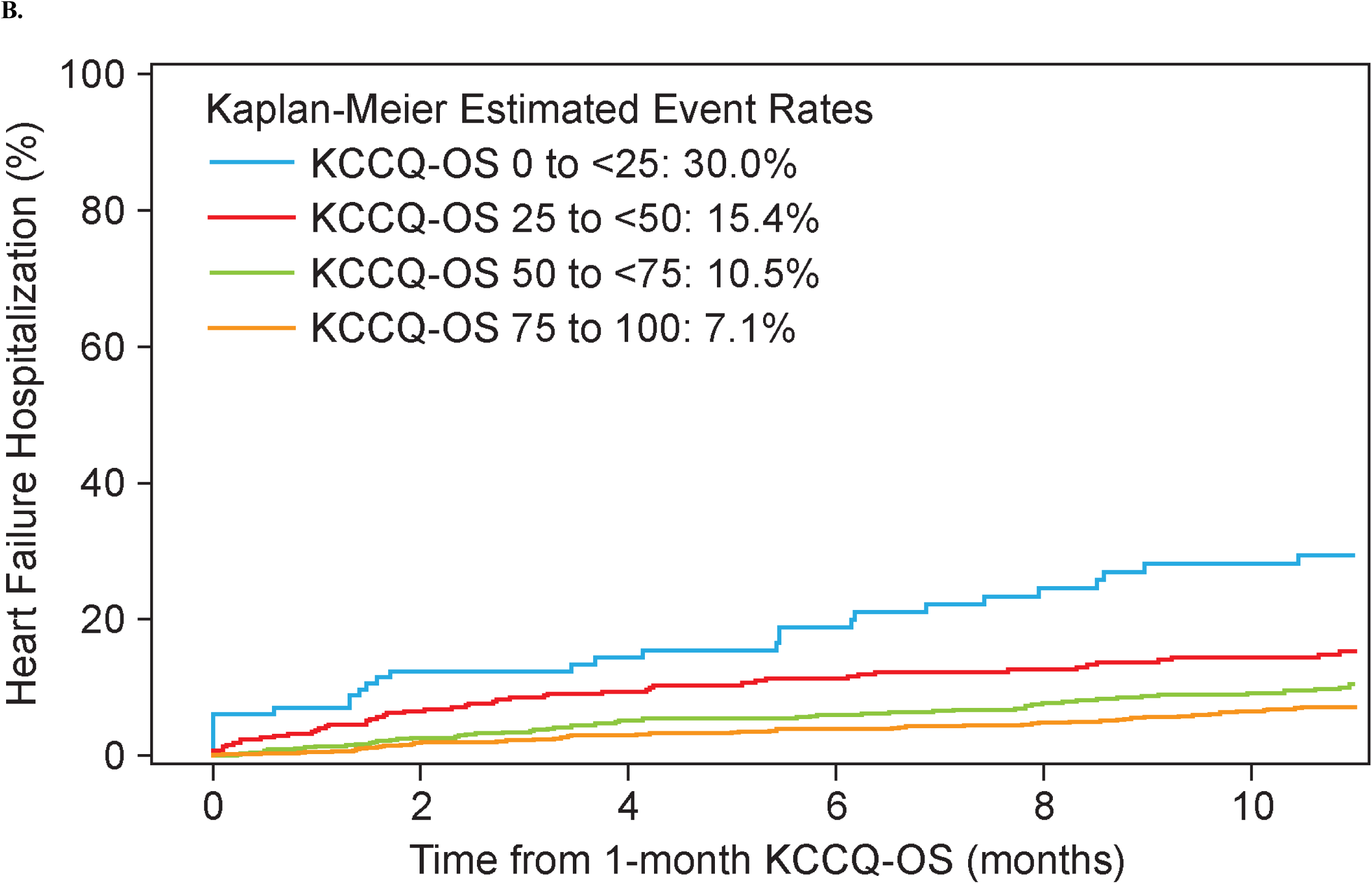
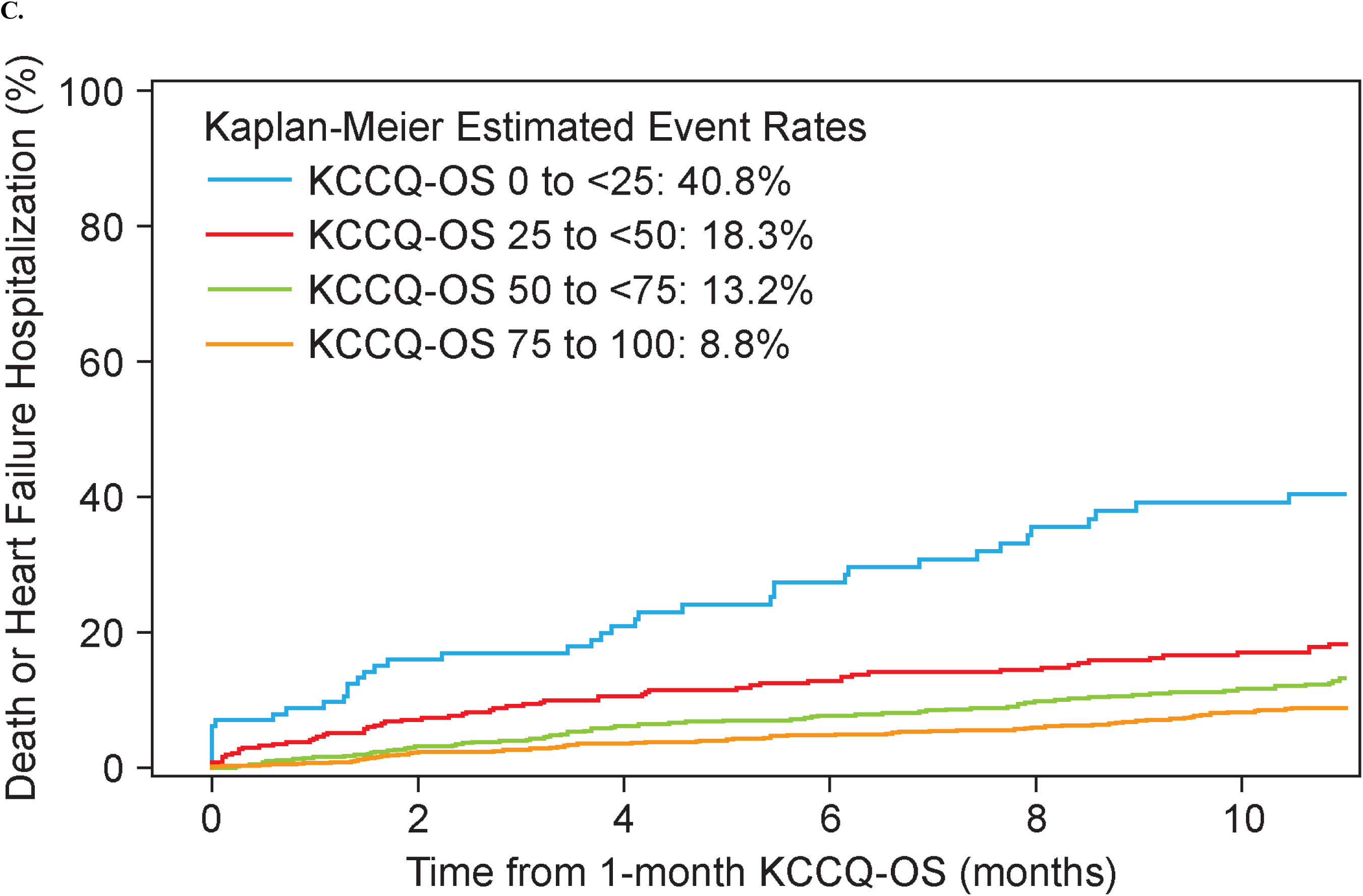
Kaplan-Meier event curves according to 1-month scores on the KCCQ Overall Summary Scale. A. Death. B. Heart failure hospitalization. C. Composite of death or heart failure hospitalization. KCCQ, Kansas City Cardiomyopathy Questionnaire. KCCQ-OS <25: n=133, KCCQ-OS 25 to <50: n=444, KCCQ-OS 50 to <75: n=781, KCCQ-OS 75 to 100: n=987

## Data Availability

Research data are not shared

## Data Sharing

Research data are not shared.

## Disclaimer

This article reflects the views of the authors and does not necessarily represent the practices, policies, requirements, or recommendations of the U.S. Food and Drug Administration. The mention of commercial products, their sources, or their use in connection with material reported here is not to be construed as either an actual or implied endorsement of such products by the Department of Health and Human Services.

