## Supplemental Tables and Figures for "Validation of the Kansas City Cardiomyopathy Questionnaire in Patients with Tricuspid Regurgitation: The Tri-QOL Study"

**Supplemental Table 1. Summary of Trials Included**

| Registration # | Trial name | Device | Sponsor |
|---|---|---|---|
| | **Single-arm trials** | | |
| NCT03382457 | Clinical Study of Edwards Cardioband Tricuspid Valve Reconstruction System[1] | Cardioband | Edwards |
| NCT03745313 | Edwards PASCAL TrAnScatheter Valve RePair System in Tricuspid Regurgitation (CLASP TR) Early Feasibility Study[2] | PASCAL | Edwards |
| NCT02471807 | Early Feasibility Study of the Edwards FORMA Tricuspid Transcatheter Repair[3] | FORMA | Edwards |
| NCT02787408 | The SPACER Trial - Repair of Tricuspid Valve Regurgitation Using the Edwards TricuSPid TrAnsCatheter REpaiR System | FORMA | Edwards |
| NCT04614402 | Transcatheter Repair of Tricuspid Regurgitation With Edwards PASCAL Transcatheter Valve Repair System (TriCLASP)[4] | PASCAL | Edwards |
| NCT04221490 | Edwards EVOQUE Tricuspid Valve Replacement: Investigation of Safety and Clinical Efficacy After Replacement of Tricuspid Valve With Transcatheter Device (TRISCEND)[5] | EVOQUE | Edwards |
| NCT04483089 | An Observational Real-world Study Evaluating Severe Tricuspid Regurgitation Patients Treated With the Abbott TriClip Device (bRIGHT)[6] | TriClip | Abbott |
| NCT03227757 | Trial to Evaluate Treatment With Abbott Transcatheter Clip Repair System in Patients With Moderate or Greater Tricuspid Regurgitation (TRILUMINATE)[7] | TriClip | Abbott |
| | **Randomized trials** | | |
| NCT04097145 | Edwards PASCAL Transcatheter Valve Repair System Pivotal Clinical Trial (CLASP II TR) | PASCAL | Edwards |
| NCT04482062 | TRISCEND II Pivotal trial | EVOQUE | Edwards |
| NCT03904147 | TRILUMINATE Pivotal trial[8] | TriClip | Abbott |

**Supplemental Table 2. Study Design**

| Psychometric Property | Patients | N | Reference Measure(s) | Analysis |
|---|---|---|---|---|
| Internal Consistency | Patients at baseline | 2645 | Baseline KCCQ items | Cronbach's alpha |
| Test-Retest Reproducibility | Patients clinically stable (1 to 6 months) | 803 | Change in KCCQ scores | Intraclass correlation |
| Responsiveness | TTVI patients at 1 month | 1323 | 1-month change in KCCQ scores | Cohen's D effect size |
| Criterion Validity (Score of Interest) | | | | |
| Physical Limitations Score | Patients at 1 month | 1255 (6MWD), 2220 (NYHA), 1553 (PCS) | 6MWD, NYHA class, SF-12 PCS | Spearman correlation |
| Symptom Scale Score | Patients at 1 month | 2278 | NYHA class | Spearman correlation |
| Quality of Life Score | Patients at 1 month | 1603 | SF-12 MCS | Spearman correlation |
| Social Limitation Score | Patients at 1 month | 1486 | SF-12 MCS | Spearman correlation |
| Clinical Summary Score | Patients at 1 month | 2279 | NYHA class | Spearman correlation |
| Overall Summary Score | Patients at 1 month | 2279 | NYHA class | Spearman correlation |
| Change in KCCQ Domain and Summary Scores | Patients at 1 month | 2326 | Change in NYHA class | Linear trend test |
| Change in KCCQ-Overall Summary Score | TTVI patients (non-FORMA) at 1 month | 1199 | Change in TR grade | Linear trend test, multivariable linear regression |
| Prognostic Implications | Patients at 1 month | 2345 | Death or heart failure hospitalization (1 month to 1 year) | Cox proportional hazards regression |

KCCQ, Kansas City Cardiomyopathy Questionnaire; TTVI, transcatheter tricuspid valve intervention; 6MWD, 6-minute walk distance; NYHA New York Heart Association; PCS, physical components summary score; MCS, mental components summary score; TR, tricuspid regurgitation

**Supplemental Table 3. Baseline Characteristics of Study Cohort, Stratified by Sex**

| | Men n=1035 | Women n=1658 |
|---|---|---|
| Enrolled in United States | 56.0% | 66.7% |
| Enrolled in randomized trial | 39.2% | 46.4% |
| Age, years | 78.6 ± 8.2 | 78.6 ± 7.9 |
| Body mass index, $kg/m^2$ | 26.4 ± 4.4 | 26.7 ± 6.2 |
| Hypertension | 81.9% | 85.5% |
| Pulmonary hypertension | 60.0% | 64.9% |
| Chronic lung disease | 17.1% | 14.0% |
| Atrial fibrillation/flutter | 92.0% | 91.2% |
| Coronary artery disease | 22.3% | 15.3% |
| Prior myocardial infarction | 14.2% | 7.1% |
| Prior percutaneous coronary intervention | 28.0% | 13.7% |
| Prior coronary bypass graft surgery | 25.2% | 9.8% |
| Prior aortic valve surgery | 15.3% | 16.1% |
| Prior mitral valve surgery | 19.2% | 26.1% |
| Pacemaker or implantable defibrillator | 28.5% | 24.2% |
| Prior stroke | 9.6% | 10.3% |
| Heart failure hospitalization in the past 12 months | 40.9% | 37.0% |
| Tricuspid regurgitation | | |
| None/trace/mild | 0.4% | 0.3% |
| Moderate | 5.3% | 6.0% |
| Severe | 31.9% | 35.2% |
| Massive | 27.3% | 30.3% |
| Torrential | 35.1% | 28.2% |
| Left ventricular ejection fraction, % | 53.7 ± 10.6 | 57.1 ± 9.9 |
| TAPSE, cm | 1.6 ± 0.4 | 1.6 ± 0.4 |
| Right ventricular FAC, % | 36.2 ± 7.9 | 38.8 ± 8.0 |
| Kansas City Cardiomyopathy Questionnaire scores | | |
| Physical Limitations | 57.8 ± 25.5 | 50.3 ± 25.0 |
| Symptoms | 58.3 ± 25.1 | 52.1 ± 24.8 |
| Quality of Life | 48.3 ± 25.4 | 43.1 ± 23.8 |
| Social Limitations | 51.4 ± 30.6 | 46.4 ± 29.7 |
| Clinical Summary | 58.0 ± 23.1 | 51.2 ± 22.7 |
| Overall Summary | 54.0 ± 23.1 | 48.1 ± 22.4 |
| NYHA class | | |
| 1 | 1.0% | 0.4% |
| 2 | 32.1% | 26.2% |
| 3 | 61.3% | 68.4% |
| 4 | 5.6% | 5.0% |

TAPSE, tricuspid annular plane systolic exertion (normal >1.7 cm); FAC, fractional area of change (normal >35%); KCCQ, Kansas City Cardiomyopathy Questionnaire

**Supplemental Table 4. Internal Consistency, Test-Retest Reproducibility, and Responsiveness of the KCCQ, Stratified by Sex**

| Property | Consistency | Test-Retest Reproducibility | | Responsiveness | |
|---|---|---|---|---|---|
| **Analytic Cohort** | **All patients** | **Stable patients** | | **TTVI patients** | |
| **Time Frame** | **Baseline** | **1 to 6 Months** | | **Baseline to 1 Month** | |
| **Test** | **Cronbach's Alpha** | **Mean Difference (95% CI)** | **ICC** | **Mean Difference (95% CI)** | **Cohen's D** |
| **Men** | **n=1010** | **n=271** | | **n=543** | |
| Physical limitation | 0.84 | 0.7 (-1.8 to 3.2) | 0.64 | 8.8 (6.8 to 10.7) | 0.34 |
| Symptoms | 0.83 | 1.1 (-1.3 to 3.5) | 0.55 | 15.0 (13.0 to 17.0) | 0.60 |
| Quality of life | 0.79 | 1.9 (-0.5 to 4.4) | 0.63 | 18.2 (16.0 to 20.4) | 0.73 |
| Social limitation | 0.79 | 0.6 (-2.7 to 3.9) | 0.59 | 14.1 (11.5 to 16.6) | 0.47 |
| Clinical summary | NA | 1.1 (-1.1 to 3.2) | 0.61 | 11.9 (10.1 to 13.7) | 0.51 |
| Overall summary | NA | 1.0 (-1.2 to 3.2) | 0.64 | 13.9 (12.1 to 15.8) | 0.61 |
| **Women** | **n=1635** | **n=600** | | **n=780** | |
| Physical limitation | 0.83 | -0.5 (-2.2 to 1.1) | 0.69 | 14.4 (12.7 to 16.2) | 0.57 |
| Symptoms | 0.83 | 0.2 (-1.3 to 1.6) | 0.69 | 19.8 (18.1 to 21.4) | 0.80 |
| Quality of life | 0.75 | 1.9 (0.0 to 3.7) | 0.63 | 23.6 (21.8 to 25.5) | 0.99 |
| Social limitation | 0.78 | 0.9 (-1.2 to 3.0) | 0.66 | 18.3 (16.0 to 20.6) | 0.63 |
| Clinical summary | NA | 0.0 (-1.3 to 1.3) | 0.73 | 17.3 (15.8 to 18.7) | 0.76 |
| Overall summary | NA | 0.8 (-0.5 to 2.1) | 0.74 | 19.2 (17.7 to 20.7) | 0.86 |

KCCQ, Kansas City Cardiomyopathy Questionnaire; TTVI, transcatheter tricuspid valve intervention; Cronbach's Alpha ($\alpha \geq 0.9$ indicates excellent consistency [but may also indicate redundancy]; $0.9 > \alpha \geq 0.8$ is good; $0.8 > \alpha \geq 0.7$ is acceptable; $0.7 > \alpha \geq 0.6$ is questionable; $\alpha < 0.6$ is poor); ICC, intra-class correlation (agreement: 0-0.2 poor, 0.3-0.4 fair, 0.5-0.6 moderate, 0.7-0.8 strong, >0.8 excellent); Coehn's D (0.2 to 0.3 small effect; ~0.5 medium effect; ≥0.8 large effect)

**Supplemental Table 5. Spearman Correlations of KCCQ Domain and Summary Score with Other Measures, Stratified by Sex**

| **Men** | **NYHA** | **SF-12 PCS** | **SF-12 MCS** | **6MWD** |
|---|---|---|---|---|
| Physical limitation | -0.54 (-0.58 to -0.49) | 0.72 (0.67 to 0.76) | | 0.47 (0.40 to 0.54) |
| Symptoms | -0.52 (-0.56 to -0.47) | | | |
| Quality of life | | | 0.59 (0.53 to 0.64) | |
| Social limitation | | | 0.56 (0.50 to 0.62) | |
| Clinical summary | -0.57 (-0.61 to -0.52) | | | |
| Overall summary | -0.55 (-0.59 to -0.50) | | | |

| **Women** | **NYHA** | **SF-12 PCS** | **SF-12 MCS** | **6MWD** |
|---|---|---|---|---|
| Physical limitation | -0.43 (-0.47 to -0.38) | 0.67 (0.63 to 0.70) | | 0.51 (0.46 to 0.56) |
| Symptoms | -0.49 (-0.53 to -0.45) | | | |
| Quality of life | | | 0.58 (0.54 to 0.62) | |
| Social limitation | | | 0.53 (0.48 to 0.58) | |
| Clinical summary | -0.50 (-0.54 to -0.46) | | | |
| Overall summary | -0.50 (-0.54 to -0.46) | | | |

All $p<0.001$

KCCQ, Kansas City Cardiomyopathy Questionnaire; NYHA, New York Heart Association; SF-12 PCS, Medical Outcomes Study Short-Form 12 Physical Components Summary score; SF-12 MCS, Medical Outcomes Study Short-Form 12 Mental Components Summary score; 6MWD, 6-minute walk distance

**Supplemental Table 6. Mean Change in KCCQ Scores According to Change in NYHA Class from Baseline to 1 Month, Stratified by Sex**

| Men | Change in NYHA Class from Baseline to 1 Month | | | | |
|---|---|---|---|---|---|
| | **Improvement by 2 or 3 n=110** | **Improvement by 1 n=382** | **No change n=348** | **Worsening n=37** | **p-value for trend** |
| Physical limitation | 17.2 ± 22.4 | 11.0 ± 22.2 | 1.5 ± 20.6 | -6.3 ± 21.0 | 0.001 |
| Symptoms | 27.4 ± 22.8 | 15.6 ± 23.1 | 5.0 ± 22.7 | 1.1 ± 21.4 | <0.001 |
| Quality of life | 32.6 ± 24.2 | 19.8 ± 25.3 | 10.1 ± 24.3 | -1.2 ± 25.1 | <0.001 |
| Social limitation | 28.5 ± 32.1 | 14.4 ± 27.9 | 5.7 ± 26.9 | -6.2 ± 25.2 | 0.003 |
| Clinical summary | 22.2 ± 20.1 | 13.4 ± 20.3 | 3.2 ± 19.4 | -1.3 ± 18.0 | <0.001 |
| Overall summary | 26.0 ± 20.2 | 15.3 ± 20.8 | 5.5 ± 19.7 | -1.9 ± 20.0 | <0.001 |

| **Women** | **Improvement by 2 or 3 n=222** | **Improvement by 1 n=645** | **No change n=479** | **Worsening n=65** | **p-value for trend** |
|---|---|---|---|---|---|
| Physical limitation | 21.5 ± 25.6 | 15.5 ± 23.1 | 8.0 ± 21.9 | -6.0 ± 22.3 | <0.001 |
| Symptoms | 30.3 ± 23.4 | 19.5 ± 21.9 | 11.0 ± 21.6 | 0.8 ± 26.2 | <0.001 |
| Quality of life | 34.6 ± 27.4 | 24.5 ± 25.1 | 15.0 ± 25.1 | -1.1 ± 21.4 | <0.001 |
| Social limitation | 30.8 ± 30.8 | 18.0 ± 30.0 | 11.0 ± 29.6 | -4.6 ± 29.9 | <0.001 |
| Clinical summary | 26.0 ± 21.0 | 17.6 ± 19.4 | 9.7 ± 18.7 | -3.7 ± 22.8 | <0.001 |
| Overall summary | 29.4 ± 21.8 | 19.6 ± 20.3 | 11.5 ± 19.8 | -3.3 ± 20.9 | <0.001 |

KCCQ, Kansas City Cardiomyopathy Questionnaire; NYHA, New York Heart Association

**Supplemental Table 7. Mean Change in KCCQ Score According to Change in TR Grade at 1 Month after TTVI, Stratified by Sex**

| | Unadjusted Change in TR Grade from Baseline to 1 Month | | | | | | | |
|---|---|---|---|---|---|---|---|---|
| **Men** | **-4 n=36** | **-3 n=111** | **-2 n=151** | **-1 n=117** | **0 n=52** | **p-value** | **Estimate (95% CI) for change in KCCQ per 1-grade reduction in TR*** | **p-value** |
| Physical limitation | 10.1 ± 23.9 | 12.8 ± 24.7 | 7.1 ± 22.9 | 6.0 ± 23.1 | 4.1 ± 19.6 | 0.076 | 1.7 (-0.5 to 3.9) | 0.124 |
| Symptoms | 19.0 ± 26.0 | 16.9 ± 25.8 | 17.4 ± 22.6 | 11.7 ± 22.4 | 7.6 ± 22.3 | 0.009 | 2.7 (0.7 to 4.6) | 0.007 |
| Quality of life | 21.9 ± 31.3 | 20.6 ± 27.5 | 18.8 ± 25.8 | 17.7 ± 24.6 | 10.1 ± 22.1 | 0.026 | 2.9 (0.6 to 5.1) | 0.014 |
| Social limitation | 23.7 ± 31.2 | 15.7 ± 28.8 | 13.9 ± 29.0 | 10.8 ± 28.5 | 8.0 ± 24.8 | 0.008 | 2.1 (-0.6 to 4.7) | 0.132 |
| Clinical summary | 14.6 ± 22.9 | 14.8 ± 22.9 | 12.4 ± 19.8 | 8.9 ± 20.2 | 5.7 ± 17.6 | 0.012 | 2.3 (0.4 to 4.2) | 0.018 |
| Overall summary | 17.9 ± 26.0 | 16.6 ± 22.8 | 14.5 ± 20.6 | 11.9 ± 20.4 | 7.0 ± 18.2 | 0.006 | 2.3 (0.3 to 4.2) | 0.023 |
| **Women** | **n=72** | **n=178** | **n=258** | **n=145** | **n=54** | **p-value** | **Estimate (95% CI)** | **p-value** |
| Physical limitation | 14.8 ± 24.2 | 17.9 ± 23.4 | 14.5 ± 23.9 | 10.9 ± 25.5 | 12.5 ± 18.0 | 0.209 | 1.0 (-0.9 to 2.9) | 0.318 |
| Symptoms | 21.6 ± 24.4 | 23.9 ± 21.6 | 18.5 ± 22.0 | 14.8 ± 23.7 | 14.6 ± 21.0 | 0.007 | 2.9 (1.2 to 4.5) | <0.001 |
| Quality of life | 25.9 ± 29.3 | 27.8 ± 24.6 | 25.2 ± 25.5 | 18.1 ± 25.9 | 15.4 ± 24.9 | 0.001 | 3.4 (1.5 to 5.4) | <0.001 |
| Social limitation | 23.0 ± 32.9 | 23.4 ± 31.5 | 17.8 ± 29.7 | 12.9 ± 28.7 | 12.2 ± 26.7 | 0.007 | 3.1 (0.7 to 5.6) | 0.011 |
| Clinical summary | 18.5 ± 22.5 | 21.3 ± 19.3 | 16.7 ± 20.0 | 12.9 ± 20.8 | 13.6 ± 16.0 | 0.018 | 2.0 (0.5 to 3.6) | 0.011 |
| Overall summary | 21.6 ± 24.1 | 23.2 ± 19.9 | 19.2 ± 20.8 | 14.7 ± 21.0 | 13.6 ± 18.0 | 0.002 | 2.7 (1.1 to 4.3) | 0.001 |

KCCQ, Kansas City Cardiomyopathy Questionnaire; TR, tricuspid regurgitation; TTVI, transcatheter tricuspid valve intervention

**Supplemental Table 8. Association of KCCQ-OS Score with Death and Heart Failure Hospitalization, Stratified by Sex**

| | Association of KCCQ-OS with Endpoint[a] (per 10-point decrement) | | | | Association of Change in KCCQ-OS with Endpoint[b] (per 10-point increase) | | | |
|---|---|---|---|---|---|---|---|---|
| **Men** | **Unadjusted HR (95% CI)** | **p-value** | **Adjusted HR[3] (95% CI)** | **p-value** | **Unadjusted HR[4] (95% CI)** | **p-value** | **Adjusted HR[5] (95% CI)** | **p-value** |
| Death | 1.30 (1.16-1.46) | <0.001 | 1.31 (1.16-1.47) | <0.001 | 0.80 (0.68-0.94) | 0.007 | 0.79 (0.66-0.94) | 0.007 |
| HFH | 1.26 (1.16-1.36) | <0.001 | 1.24 (1.14-1.35) | <0.001 | 0.83 (0.73-0.95) | 0.008 | 0.83 (0.72-0.95) | 0.008 |
| Death or HFH | 1.28 (1.19-1.37) | <0.001 | 1.27 (1.18-1.36) | <0.001 | 0.82 (0.74-0.91) | <0.001 | 0.81 (0.73-0.91) | <0.001 |
| **Women** | **Unadjusted HR (95% CI)** | **p-value** | **Adjusted HR[c] (95% CI)** | **p-value** | **Unadjusted HR[d] (95% CI)** | **p-value** | **Adjusted HR[e] (95% CI)** | **p-value** |
| Death | 1.37 (1.17-1.60) | <0.001 | 1.41 (1.20-1.66) | <0.001 | 0.80 (0.63-1.00) | 0.050 | 0.80 (0.63-1.02) | 0.070 |
| HFH | 1.25 (1.15-1.36) | <0.001 | 1.24 (1.14-1.35) | <0.001 | 0.76 (0.67-0.87) | <0.001 | 0.77 (0.68-0.89) | <0.001 |
| Death or HFH | 1.26 (1.17-1.36) | <0.001 | 1.25 (1.16-1.35) | <0.001 | 0.78 (0.69-0.88) | <0.001 | 0.79 (0.70-0.89) | <0.001 |

KCCQ-OS, Kansas City Cardiomyopathy Questionnaire-overall summary score; HR, hazard ratio; CI, confidence interval; HFH, heart failure hospitalization

[a] KCCQ-OS at 1 month from trial enrollment, with endpoints assessed 1 month through 1 year of follow-up.

[b] Change in KCCQ-OS from baseline to 1 month from TTVI, with endpoints assessed 1 month through 1 year of follow-up. Includes only patients in single-arm trials who were treated with TTVI.

[c] Adjusted for age, sex, body mass index, chronic lung disease, atrial fibrillation/flutter, coronary artery disease, prior myoc ardial infarction, prior coronary artery bypass graft surgery, prior stroke, permanent pacemaker, and left ventricular ejection fra ction

[d] Adjusted for baseline KCCQ-OS

[e] Adjusted for baseline KCCQ-OS, age, sex, body mass index, chronic lung disease, atrial fibrillation/flutter, coronary artery disease, prior myocardial infarction, prior coronary artery bypass graft surgery, prior stroke, permanent pacemaker, and left ventricular ejection fraction
